# Unmasking True Clinical Competence: The Importance of Adaptive and Open-Ended Evaluation for Large Language Models in Cardiology

**DOI:** 10.1101/2025.05.17.25327814

**Authors:** Chieh-Ju Chao, Abhinav Kumar, Aakash Mishra, Yu-Chiang Wang, Chieh-Mei Tsai, Nima Baba Ali, Somanshu Sharma, Juan M. Farina, Reza Arsanjani, Yucheng Jiang, Monica Lam

**Affiliations:** Department of Cardiovascular Medicine, Mayo Clinic, Rochester, MN, USA; Institute of Human-centered Artificial Intelligence, Stanford University, Palo Alto, CA, USA; Department of Computer Science, Stanford University, Palo Alto, CA, USA; Department of Cardiovascular Diseases, Mayo Clinic, Scottsdale, AZ, USA

**Keywords:** Large language model, cardiology, clinical reasoning, board examination, evaluation framework, artificial intelligence

## Abstract

**Background:** Large language models (LLMs) achieve impressive accuracy on multiple-choice (MC) medical examinations, but this performance alone may not accurately reflect their clinical reasoning abilities. MC evaluations of LLMs risk inflating apparent competence by rewarding recall rather than genuine clinical adaptability, particularly in specialized medical domains such as cardiology.

**Aim:** To rigorously evaluate the clinical reasoning, contextual adaptability, and answer-reasoning consistency of a state-of-the-art reasoning-based LLM in cardiology using MC, open-ended, and clinically modified question formats.

**Methods:** We assessed GPT o1-preview using 185 board-style cardiology questions from the American College of Cardiology Self-Assessment Program (ACCSAP) in MC and open-ended formats. A subset of 66 questions underwent modifications of critical clinical parameters (e.g., ascending aorta diameter, ejection fraction) to evaluate model adaptability to context changes. The model’s answer and reasoning correctness were graded by cardiology experts. Statistical differences were analyzed using exact McNemar tests.

**Results:** GPT o1-preview demonstrated high baseline accuracy on MC questions (93.0% answers, 92.4% reasoning). Performance significantly decreased with open-ended questions (80.0% answers, 80.5% reasoning; p<0.001). For modified MC questions, accuracy decreased significantly (answers: 93.9% to 66.7%; reasoning: 93.9% to 71.2%; both p<0.001), as well as answer-reasoning concordance (93.9% to 66.7%, p<0.001).

**Conclusions:** Using existing MC question formats substantially overestimates the performance and clinical reasoning capabilities of GPT o1-preview. Incorporating open-ended, clinically adaptive questions and evaluating answer-reasoning concordance are essential for accurately assessing the real-world clinical decision-making competencies of LLMs in cardiology.

## Introduction

Recent advancements in large language models (LLMs) have shown promising performance across numerous medical tasks, with several studies suggesting these models can match or exceed human experts on standardized medical board-style examinations^1–6^. Such models include generic LLMs, medicine-specific LLMs^7,8^, and more recently, “reasoning” models, such as GPT o1-preview, which decompose a query into logical steps before arriving at a final answer ^9^. Although LLM systems have been reported to achieve comparable or suprahuman performance in these questions^1–6^, there is still a question of whether current evaluation methodologies truly reflect these models’ capability in clinical settings. Moreover, most studies have focused on general medical license exam questions ^1,10,11^, with some evaluation in subspecialties like Cardiology^12,13^.

Current evaluation frameworks for LLMs in medicine rely heavily on multiple-choice (MC) question formats ^14^. MC examinations, as being questioned for their effectiveness in evaluating human medical trainees ^15–17^, can also be misleading when assessing LLM performance. The prevalence of MC question answering frameworks in current LLM research is largely due to pragmatic considerations. MC answers are easily evaluated and can be scaled across a large number of questions and multiple datasets. MC questions can be compromised by cues in the answer choices or “best guess” strategies ^16–19^, which LLMs can exploit without necessarily understanding the underlying clinical concepts. In addition, since many board-style questions exist in widely available databases ^1,11^, there is a risk that some have been seen during an LLM’s pre-training phase, which can inflate accuracy during evaluation.

An alternative approach is to evaluate using long-form, open-ended questions ^8,20^, where the model must generate a complete written rationale and final answer without relying on multiple-choice cues. However, evaluating open-ended responses presents significant challenges, particularly in specialized medical domains, where expertise is required to judge answer correctness ^21^. In cardiology practice, critical clinical decisions hinge on key quantitative metrics, such as left ventricular ejection fraction for guiding implantable cardioverter-defibrillator therapy and ascending aorta dimensions for surgical repair thresholds. The capability of making decisions according to these contextual clinical variations would serve as a critical indicator of clinical reasoning capability in LLM assessments. This approach, which evaluates a model’s ability to interpret and adapt to shifting clinical parameters, has not been systematically incorporated into prior LLM evaluation frameworks ^1–6^.

In this study, we present an evaluation framework aimed at overcoming the limitations of the conventional MC question format and more accurately gauging LLM performance, including answering and reasoning accuracy in the field of cardiology. In doing so, we highlight both the potential and the current limitations of GPT o1-preview, a state-of-the-art reasoning LLM, in cardiology practice.

## Methods

In this work, we evaluated GPT o1-preview, one of OpenAI’s latest LLMs that has a specific emphasis on its reasoning capacity on a set of 185 subspecialty board-style cardiology questions. We use two primary question formats, including multiple-choice and open-ended, to evaluate whether performance differs in the presence (or absence) of explicit answer options. We additionally assess the model’s ability to understand the clinical context by modifying a subset of 66 questions, in which we change key clinical details so that the correct answer differs from the original correct answer. By comparing performance on original vs. modified questions using expert cardiologist evaluators, we evaluate GPT o1-preview’s ability to adapt to nuanced changes in clinical scenarios. **Figure 1** illustrates our study design.

**Figure 1.**
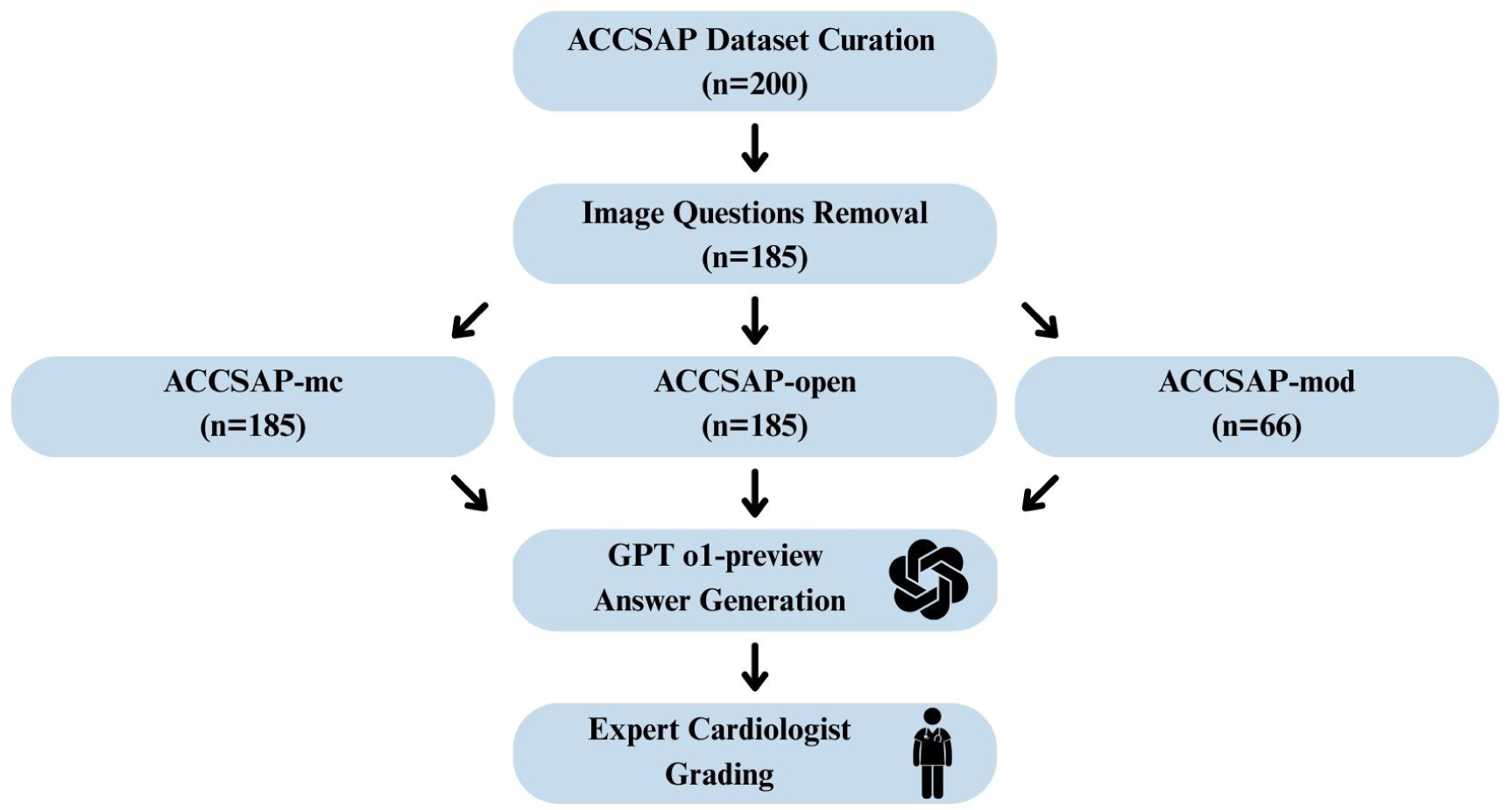
Study design for assessing clinical reasoning capabilities of GPT o1-preview. The question dataset was curated from the ACCSAP, excluding image-based questions. Questions were formulated into multiple-choice (ACCSAP-mc), open-ended (ACCSAP-open), and clinically modified (ACCSAP-mod) formats. Model responses underwent expert grading to quantify performance across question formats.

### Baseline Question Dataset

We conducted an evaluation of GPT o1-preview on a dataset of 185 board-style cardiology questions. We leveraged the American College of Cardiology’s Self-Assessment Program (ACCSAP), a board-preparation resource designed for the cardiology subspecialty board exam. This question bank, which is behind a paywall, contains a total of 640 questions covering various cardiology topics and is less likely to be leaked for LLM training due to its restricted access.

To create a representative benchmark, we initially used ACCSAP’s random selection function to obtain 200 questions, ensuring balanced coverage across all question categories. As our study focuses solely on evaluating the model’s ability to understand clinical text, we followed prior research methodologies and excluded questions that contained figures in the question stem^12^, leaving 185 questions. This allows for a more precise assessment of the model’s textual comprehension without the confounding factor of medical image interpretation.

To extend our evaluation framework beyond the provided multiple-choice questions, we introduced two question formats of the original dataset:

1. Multiple choice questions with a “None” option (ACCSAP-mc, n=185) – We added an additional “None” option to the provided answer choices to mitigate common process-of-elimination strategies used by reasoning models and simulate a more realistic clinical decision-making scenario.
2. Open-ended questions (ACCSAP-open, n=185) – We removed answer choices and revised the question stem to prompt models to generate the best possible answer in an open-ended format, encouraging more nuanced reasoning rather than pattern recognition.

### Clinical Context Question Modification

Building on the baseline ACCSAP dataset, we aimed to test GPT o1-preview’s ability to adapt to nuanced changes in clinical parameters rather than simple recall for memorized solutions. All 185 questions were reviewed for modifiable content, including specific symptoms and numerical measurements that are critical for informed clinical decision-making.

A subset of 66 questions (ACCSAP-mod) was selected for contextual modification to evaluate LLM’s flexibility in clinical reasoning^22^. Specifically, each modification involved altering one or more key clinical parameters, such as decreasing the ascending aorta diameter from 35 mm to 55 mm or adjusting the left ventricular ejection fraction (LVEF) from 15% to 55%, so that the correct answer to the modified question would differ from the original question’s correct answer. Modifications were made according to current cardiology guidelines to ensure each revised question had a legitimate new best answer. **Table 1** provided a few examples of how the questions were modified (not the questions used in our test).

**Table 1.** Examples of Question Modification.

| Original Question* | Modified Question | Change in the answer |
| --- | --- | --- |
| A 63-year-old man with a medical history of coronary artery disease, for which he underwent percutaneous coronary intervention 3 years earlier. A recent echocardiogram showed left ventricular ejection fraction (LVEF) 26%, unchanged from 6 months prior. He is optimized on guideline-directed medical therapy (GDMT).<br>An electrocardiogram shows normal sinus rhythm and left bundle branch block (LBBB) with QRS width 160 msec. What's the next step? | A 63-year-old man with a medical history of coronary artery disease, for which he underwent percutaneous coronary intervention 3 years earlier. A recent echocardiogram showed left ventricular ejection fraction (LVEF) 26%, unchanged from 6 months prior. He is optimized on guideline-directed medical therapy (GDMT). An electrocardiogram shows normal sinus rhythm and **first-degree AV block**. What's the next step? | **Original:** Implant a biventricular implantable cardioverter-defibrillator.<br>**Modified:** Implant a single-chamber implantable cardioverter-defibrillator. |
| A 58-year-old man with a history of bicuspid aortic valve (BAV). An echocardiogram 1 year earlier showed normal left ventricular (LV) function, and a BAV with a mean gradient across the aortic valve (AoV) was 16 mm Hg. The ascending aortic diameter was 4.6 cm. Repeated echocardiogram today shows similar LV function, and the mean gradient across the AoV is 18 mm Hg. The ascending aortic diameter is 5 cm.<br>What's the best next step? | A 58-year-old man with a history of bicuspid aortic valve (BAV). An echocardiogram 1 year earlier showed normal left ventricular (LV) function, and a BAV with a mean gradient across the aortic valve (AoV) was 16 mm Hg. The ascending aortic diameter was 4.6 cm. Repeated echocardiogram today shows similar LV function, and the mean gradient across the AoV is 18 mm Hg. **The ascending aortic diameter is 4.6 cm.**<br>What's the best next step? | **Original:** Refer to surgical intervention<br>**Modified:** Repeat echocardiogram in 1 year. |
| A 65-year-old woman with nonischemic dilated cardiomyopathy (left ventricular ejection fraction [EF] 35%) is here for follow-up. Her medications are carvedilol 6.25 mg twice daily and losartan 50 mg daily.<br>Physical examination showed normal blood pressure 137/84 and heart rate 60. Laboratory results showed normal creatinine and electrolytes.<br>What's the next best step in managing heart failure? | A 65-year-old woman with nonischemic dilated cardiomyopathy (left ventricular ejection fraction [EF] 35%) is here for follow-up. Her medications are carvedilol **25** mg twice daily and losartan 50 mg daily.<br>Physical examination showed normal blood pressure 137/84 and heart rate **35**. Laboratory results showed normal creatinine and electrolytes.<br>What's the next best step in managing heart failure? | **Original:** Substitute sacubitril/valsartan for losartan.<br>**Modified:** Reduce carvedilol. |
| A 68-year-old man presents to the clinic for a postoperative reassessment. He had a 26 mm bioprosthetic mitral valve (MV) replacement for severe mitral regurgitation 6 months earlier.<br>What's the best medication regimen for the patient? | A 68-year-old man presents to the clinic for a postoperative reassessment. He had a 26 mm bioprosthetic mitral valve (MV) replacement for severe mitral regurgitation **2 months** earlier.<br>What's the best medication regimen for the patient? | **Original:** Aspirin 81 mg daily.<br>**Modified:** Warfarin with target INR 2.5. |

| Table 1. Examples of Question Modification |  |  |
| --- | --- | --- |
| A 72-year-old man with a history of aortic stenosis (AS), hypertension, and hyperlipidemia. His medications are amlodipine 10 mg daily and atorvastatin 40 mg daily.<br>Vital signs are within normal limits. Transthoracic echocardiogram (TTE) reveals left ventricular (LV) ejection fraction 65% and mild LV hypertrophy. The aortic valve is calcified with peak velocity 4.3 m/sec, mean gradient 42 mm Hg, and calculated aortic valve area (AVA) 0.8 cm<sup>2</sup>.<br>What is the best next step? | A 72-year-old man with a history of aortic stenosis (AS), hypertension, and hyperlipidemia. His medications are amlodipine 10 mg daily and atorvastatin 40 mg daily.<br>Vital signs are within normal limits. Transthoracic echocardiogram (TTE) reveals left ventricular (LV) ejection fraction 45% and mild LV hypertrophy. **The aortic valve is calcified with peak velocity 3.2 m/sec, mean gradient 28 mm Hg,** and calculated aortic valve area (AVA) 0.8 cm<sup>2</sup>.<br>What is the best next step? | **Original:** Exercise treadmill testing.<br>**Modified:** Dobutamine stress echocardiography. |
| *Abbreviated content to demonstrate the clinical context of the original ACC questions. |  |  |

In the multiple-choice (MC) format, these modifications are designed to admit one correct answer for each question. Similarly, in the open-ended format, the question stem was reworded to incorporate the new clinical values, so that the LLM needed to derive a different final answer and reasoning. This approach distinguished truly context-aware reasoning from mere pattern matching or recall of prior training data.

For analysis, we compared the model’s performance on the original vs. modified versions of the same question in both multiple-choice and open-ended formats.

### Model Selection

We selected GPT o1-preview^23^ for this study for its advanced reasoning architecture; it goes beyond simple pattern recognition to systematically work through multi-step reasoning. Compared to earlier GPT models, GPT o1-preview uses an internal chain-of-thought approach, allowing it to “think aloud” over relevant details and integrate guidelines before arriving at a final conclusion. This design more closely mirrors the reasoning process of human experts, making o1-preview suitable for complex, context-dependent questions such as those found in advanced board-style exams. The evaluation of GPT o1-preview is performed using Microsoft Azure services, ensuring that our test data is not leaked to OpenAI for training purposes.

### Model Inference and Data Collection

For each question and each format, o1-preview was prompted with the question and asked to “answer a general practitioner’s question” by generating a final answer to the question and providing a detailed reasoning to justify the answer. In the multiple-choice question format, GPT o1-preview selected one of the provided lettered answers and then produced a rationale in free-text format. In the open-ended format, it produced a written answer and its reasoning in a free text format. See **Supplementary Section A.1** and **Supplementary Section A.2** for the specific prompts used in generating answers and reasoning.

### Evaluation

Human expert grading was first performed by YCW and CMT and reviewed by a board-certified cardiologist (CJC). Raters examined GPT-o1-preview’s final answer and accompanying reasoning beside the reference solution and assigned two independent binary labels: answer correct/incorrect and reasoning correct/incorrect. An answer was deemed correct if it exactly matched, or represented an alternative yet clinically acceptable formulation of, the ground-truth response.

The current gold standard for evaluating open-ended and reasoning-based questions relies on human clinical experts. However, this approach is often limited by availability and is not easily scalable. Therefore, we also leveraged LLM-based evaluators to assess o1-preview’s performance and compared their agreement with human evaluators^20^. See **Supplementary Section B** for implementation details. For both metrics, we evaluated the agreement between GPT-4o automated grading and the expert cardiologist grading using Cohen’s kappa (See **Supplementary Section C**).

### Statistical Analysis

We assessed question-answering performance using answer accuracy, the number of questions with the correct answer generated, reasoning accuracy, and the number of questions with the correct reasoning generated. To assess the model’s internal answer–reasoning concordance, we constructed a 2 × 2 contingency table for every question set, cross-classifying answer correctness against reasoning correctness. We defined the full-concordance rate (FCR) as the proportion of questions in which both answer and reasoning were correct (A+/R+) and the full-discordance rate (FDR) as the proportion in which both were incorrect (A–/R–).

Paired comparisons were performed with exact McNemar tests: (i) baseline multiple-choice vs. baseline open-ended; (ii) baseline vs. modified multiple-choice; and (iii) baseline vs. modified open-ended. All analyses were two-sided, with statistical significance set at *p* < 0.05, and were conducted in Python 3.11 using the *statsmodels* package (v0.14).

## Results

### Characteristics of Question Sets

A total of 185 original questions were evaluated. **Table 2** summarizes the distribution across 10 categories, with the largest subset addressing Heart Failure and Cardiomyopathies (24.3%, n=45). Each question was presented in both open-ended and multiple-choice formats for GPT o1-preview baseline evaluation. A subset of 66 questions was modified to evaluate contextual adaptation. **Table 3** describes the distribution of the modified questions by category.

**Table 2:** Distribution of ACCSAP Questions.

| Category | Number of Questions | Percentage (%) |
| --- | --- | --- |
| Heart Failure & Cardiomyopathies | 45 | 24.3 |
| Coronary Artery Disease | 26 | 14.1 |
| Valvular Heart Disease | 24 | 13.0 |
| Arrhythmias | 21 | 11.1 |
| Vascular Disease | 20 | 10.8 |
| Congenital Heart Disease | 14 | 7.6 |
| Hypertension | 13 | 7.0 |
| Pericardial Disease | 13 | 7.0 |
| Miscellaneous Topics in Cardiovascular Medicine | 7 | 3.8 |
| Systemic Disorders Affecting Circulatory System | 2 | 1.1 |
| Total | 185 | 100.0 |

**Table 3:** Distribution of Modified ACCSAP Question Subset.

| Category | Number of Questions | Percentage (%) |
| --- | --- | --- |
| Heart Failure & Cardiomyopathies | 15 | 22.7 |
| Coronary Artery Disease | 9 | 13.6 |
| Valvular Heart Disease | 9 | 13.6 |
| Vascular Disease | 9 | 13.6 |
| Arrhythmias | 7 | 10.6 |
| Hypertension | 6 | 9.1 |
| Pericardial Disease | 6 | 9.1 |
| Congenital Heart Disease | 4 | 6.1 |
| Systemic Disorders Affecting Circulatory System | 1 | 1.5 |
| Total | 66 | 100.0 |

### Baseline Question Performance

For multiple-choice questions, GPT o1-preview selected the correct answer in 93.0% of cases (n = 172), and expert cardiologists rated 92.4% of the generated reasonings as correct (n = 171). 0.5% (n=1) of multiple-choice reasonings were labeled as “alternative, clinically acceptable” solutions. Notably, the Arrhythmias, Systemic Disorders Affecting the Circulatory System, and Valvular categories all achieved 100% accuracy for both answers and reasoning. Pericardial Disease category exhibited the lowest performance, with answer and reasoning accuracies of 76.9% (n = 10) and 69.2% (n = 9), respectively.

For open-ended questions, expert evaluators independently rated 80.0% (n=145) of the answers as correct and 80.5% (n=146) of the reasonings as correct. 7.6% (n=14) of open-ended answers and 7.6% (n=14) of open-ended reasonings were rated as “alternative, clinically acceptable” solutions. Questions in the Systemic Disorders Affecting Circulatory System category had the highest open-ended answer and reasoning accuracy of 100% (n=2). Consistent with the MC format, questions in the Pericardial Disease category were the most challenging, with an answer accuracy of 69.2% (n=9) and a reasoning accuracy of 61.5% (n=8).

Overall, multiple-choice performance significantly exceeded that of the open-ended format, with answer accuracy of 93.0% versus 80.0% (p < 0.001) and reasoning accuracy of 92.4% versus 80.5% (p < 0.001). As shown in **Figure 2**, the magnitude of this gap between multiple-choice and open-ended performance varied across categories. Valvular Heart Disease had the largest difference, with answer accuracy dropping from 100.0% in multiple-choice to 75.0% in open-ended format (p=0.03).

**Figure 2.**
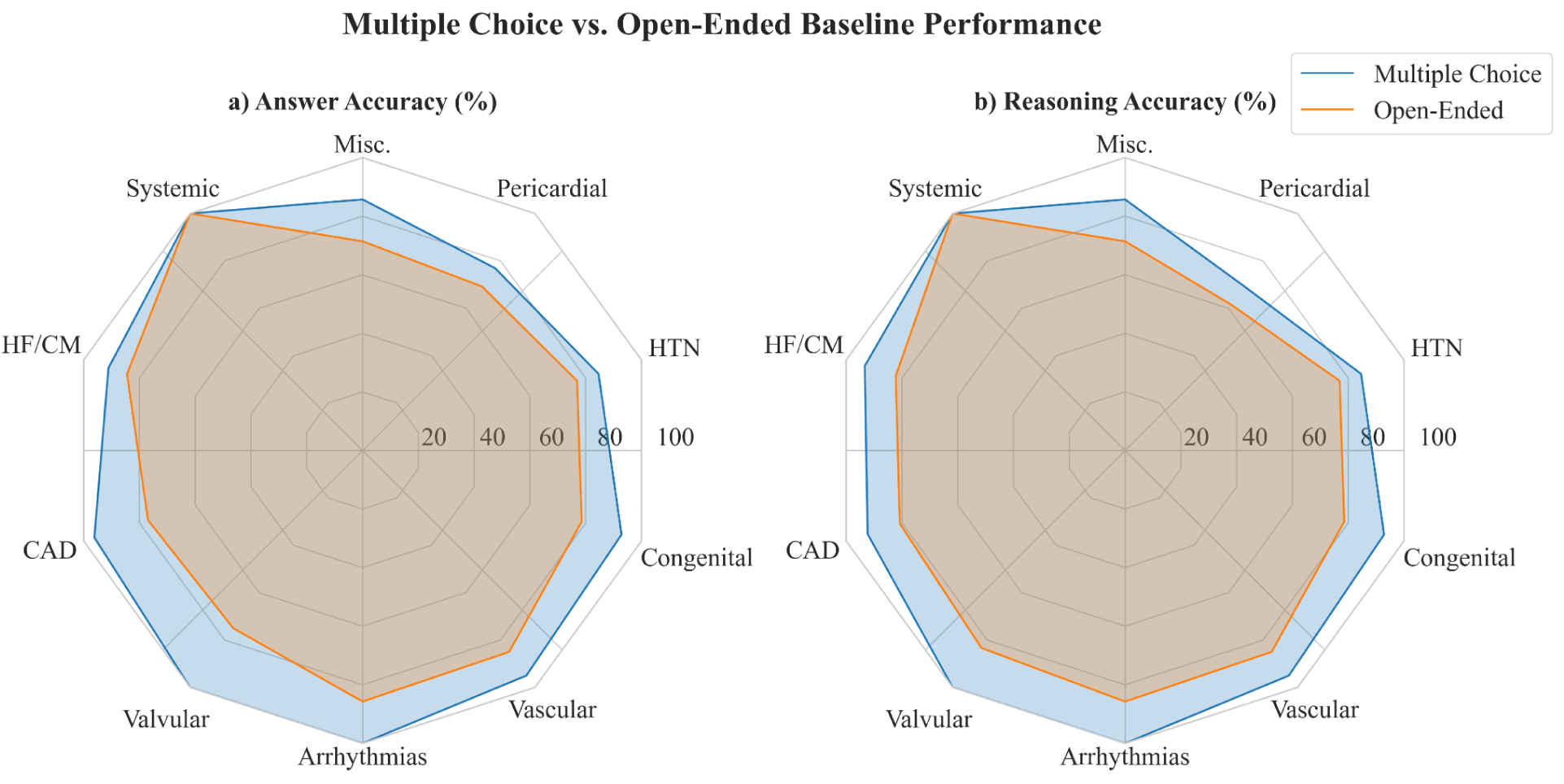
Comparison of performance between multiple-choice and open-ended formats across baseline cardiology questions. **2a** shows the percentage of correct answers, and **2b** shows the percentage of correct reasoning across each question category.

### Modified Question Performance

Baseline MC performance on the 66-question subset was strong, with overall answer and reasoning accuracies of 93.9%. After the MC questions were modified, GPT o1-preview’s answer accuracy decreased to 66.7% (p < 0.001), and reasoning accuracy dropped to 71.2% (p = 0.0015), as shown in **Figure 3**. Note, no multiple-choice reasonings were labeled as alternatively acceptable. Although no single subcategory’s difference reached significance, the aggregate differences in both answer correctness and reasoning correctness were statistically significant, suggesting that GPT o1-preview was consistently less accurate when the multiple-choice questions contained modified clinical parameters.

**Figure 3.**
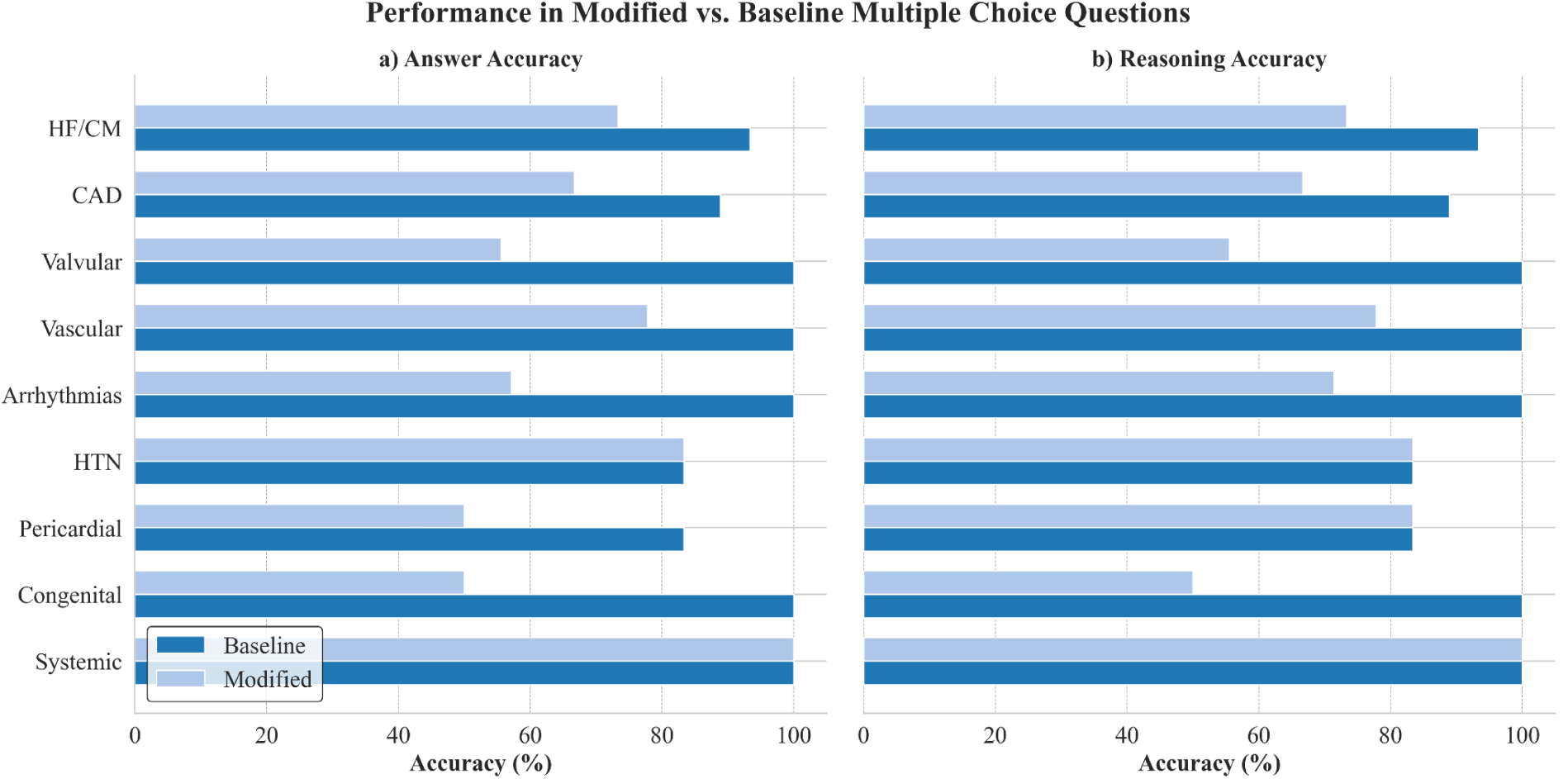
Comparison of performance between baseline and modified **multiple**-**choice** questions for each cardiology category. **3a** illustrates the percentage of correct answers, and **3b** shows the percentage of correct reasoning.

**Table 4** summarizes GPT-o1-preview’s joint answer–reasoning performance. In baseline multiple-choice questions, fully concordant outputs were observed in 170 questions (FCR = 91.9 %), whereas 6.5 % were fully discordant. When the identical items were presented in open-ended form, FCR fell to 78.9 % (n = 146) and FDR rose to 18.4 % (p < 0.001), indicating lower answer–reasoning consistency once MC answer cues were removed.

**Table 4.**
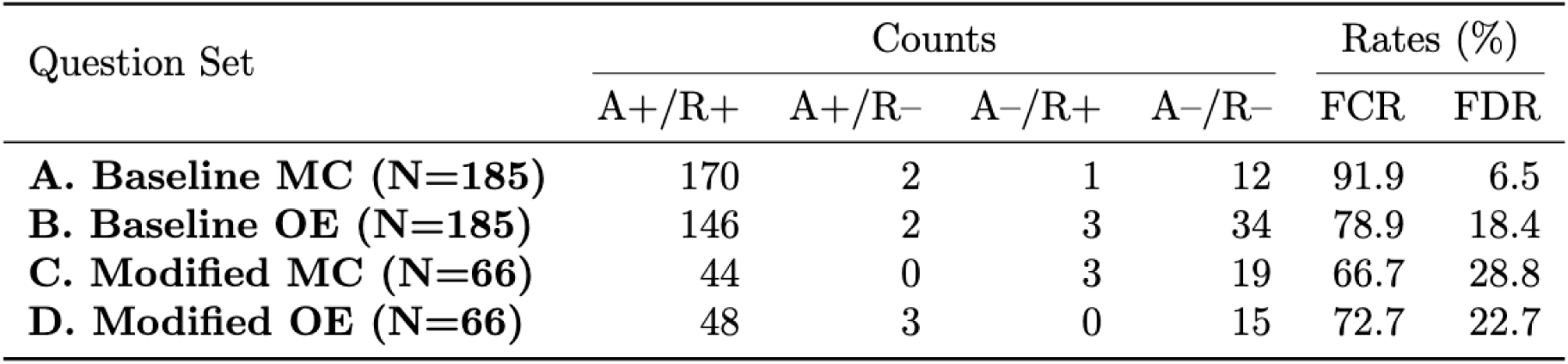
Joint answer-reasoning performance (A/R) across baseline and modified question sets. MC = multiple-choice; OE = open-ended; A+/R+ answer and reasoning both correct; A+/R– = answer correct, reasoning incorrect; A–/R+ = answer incorrect, reasoning correct; A–/R– = both incorrect. FCR = full-concordance rate (proportion of A+/R+); FDR = full-discordance rate (proportion of A–/R–). All ratings by expert cardiologists.

Among the 66 questions available in both baseline and modified versions, baseline multiple-choice performance remained highly concordant (FCR = 93.9 %) but declined to 66.7 % after clinical modifications (p < 0.001). Baseline vs. modified open-ended items showed no significant difference in concordance (77.3 % vs. 72.7 %; p = 1.0). These findings indicate that GPT-o1-preview’s performance is more vulnerable to context shifts in an MC framework than in an open-ended format.

## Discussion

The main contributions of this work include: 1) Introducing an improved evaluation framework that goes beyond conventional MC formats by incorporating open-ended questions and revised clinical contexts, enabling a more comprehensive assessment of LLMs’ clinical reasoning and answering abilities in realistic cardiology practice scenarios, and 2) Our results revealed a significant drop in LLM performance when evaluated with this framework, highlighting the limitations of current models in handling complex, clinically relevant reasoning tasks in a specialized medical specialty. Our findings challenge the conventional approach of evaluating LLMs with MC questions that may have been used in training and suggest the need for more rigorous LLM evaluation methods in the medical field that critically assess real-world clinical reasoning.

Cardiology is a specialized field with rapidly evolving evidence. Clinical decisions often depend on precise assessments like echocardiography, requiring thorough knowledge of both the evidence and clinical context for optimal outcomes^24–26^. Previous studies in this field have predominantly assessed LLM accuracy using MC questions without systematically examining the quality or consistency of the corresponding reasoning processes ^12,27–30^. Consistent with prior studies using MC questions ^12,27–30^, GPT o1-preview demonstrated high accuracy on ACCSAP-mc questions (93.0%), superseding GPT-4’s 71.0% on a different set of MC format ACCSAP questions ^12^. However, when evaluation extended beyond conventional MC format questions, o1-preview did not demonstrate the same level of capability across question formats and contextual adaptations (**Figures 2 and 3**). O1-preview’s performance significantly deteriorated when questions were reformulated into open-ended formats (ACCSAP-open, 80.0%) or even more when minimal but critical clinical context modifications were introduced (ACCSAP-mod, 66.7%). This aligns with previous findings that LLMs show reduced performance when MC questions undergo alterations in option ordering or response format ^31^.

Although the o1-preview model is claimed to possess generalized reasoning capabilities^6,32^, our findings indicate these models cannot fully replicate expert clinical decision-making in cardiology. In open-ended scenarios or contextually adapted questions, o1-preview often failed to apply logical reasoning or adapt reasoning to current clinical guidelines, similar to the inflexible reasoning failure mode described in recent studies ^22,33,34^. We observed the most significant performance drop in the valvular heart disease subcategory (from 100.0% to 75.0%), in which clinical decisions can be distinct with semantically minor changes in a single echocardiography measurement (e.g., aortic valve area 0.8 cm^2^ vs. 2.5 cm^2^). This finding suggests a limitation in detecting subtle but clinically significant contextual changes.

In some ACCSAP-mod questions, o1-preview provided reasoning corresponding to the original unmodified question despite changes in critical clinical context, implying memorization of previously encountered question-answer pairs during training rather than genuine clinical understanding. When further analyzing o1-preview’s answer–reasoning concordance, fully concordant outputs dropped from 93.9% to 66.7 % after minor clinical modifications in a multiple-choice setting, suggesting the model’s dependence on fixed answer cues rather than flexible, context-sensitive reasoning. O1’s performance was similarly low on modified MC and OE questions, indicating that the model struggled in handling “new” information in the modified clinical context, regardless of the question format. The above observations suggest that o1’s performance, to some extent, still relies on pattern recognition from seen examples instead of comprehensive understanding and reasoning based on clinical context. Prior research in human medical education also challenges the assumption that knowledge memorization equates to clinical understanding^15–17,35^. Our results, particularly the significant performance drop between baseline and modified questions, further highlight the limitations of using existing questions in assessing LLMs’ clinical capabilities. Thus, caution is warranted when interpreting and promoting results from assessments of LLM performance based on existing question sets.

As LLMs advance into complex reasoning, their clinical evaluation frameworks must also evolve. Human expert evaluation is subject to availability constraints, potential subjectivity, and limited scalability. To address these challenges, we explored the use of automated evaluation by assessing GPT-4o’s capability to judge the reasoning generated by o1-preview. While our automated evaluation using GPT-4o showed substantial agreement with expert cardiologists in assessing MC reasoning (Cohen’s kappa = 0.78), agreement was considerably lower for open-ended questions (Cohen’s kappa = 0.37 for answer correctness for baseline questions, 0.36 for modified questions). Therefore, despite being more resource-intensive, employing human experts remains the gold standard for evaluating models’ performance in open-ended answering and reasoning tasks in the medical field^36,37^.

Several limitations should be considered when interpreting our results. Although we did not include the entire ACCSAP dataset, our benchmark was randomly selected to ensure balanced representation of the original dataset while maintaining feasibility for evaluation. Excluding questions with images didn’t allow assessment of medical image interpretation; however, this design eliminates the confounding factor and is consistent with prior studies. Not all original questions were suitable for context modification due to their content. As a result, the revised questions did not cover all categories. Additionally, transitioning questions from MC to an open-ended format may introduce evaluation biases with subjective judgments. Only o1-preview was evaluated as the representative state-of-the-art reasoning model, knowing its superior performance compared to other recent models.

## Conclusion

Although LLMs achieve high accuracy on MC cardiology board questions, our study demonstrates that using established MC question benchmarks can substantially overestimate their performance, and lack assessment of their true clinical reasoning capabilities, particularly when adapting to diverse clinical contexts. Therefore, future assessment benchmarks for LLMs in cardiology should incorporate open-ended, clinically adaptive frameworks to more accurately reflect real-world clinical decision-making.

## Data Availability

The ACCSAP questions are behind a paywall but can be accessed with payment. To prevent unauthorized distribution of the revised question set, the open-ended and revised content questions are not made publicly available. However, they can be obtained upon reasonable request from the corresponding author.

## Abbreviations

MC: multiple choice
LLM: large language model
HTN: hypertension
CAD: coronary artery disease
HF/CM: heart failure & cardiomyopathies

## Supplementary Material

### Section A

#### A.1 Multiple Choice Question Answering Prompt (GPT o1-preview)

You are a cardiologist. Your task is to select the most correct answer to a general practitioner’s multiple choice question and provide the most specific and concrete reasoning. Your answer should be one of the provided multiple choice answers.

**Key Instructions**

1. Answer the question with only one best multiple choice option based on the scenario provided. Avoid choosing multiple recommendations or answers, even if more than one seems relevant.
2. Provide an explanation of reasoning for your answer under the section **Reasoning:** to justify your choice.
3. Provide your final answer under the section **Answer:**. This section should only contain the letter of the multiple choice answer you selected!

**General Tips**

Ensure your answer is a single letter (A, B, C, D, or E), corresponding to the multiple choice option that you choose. Be specific and detailed in your reasoning. If multiple options seem equally valid, choose the one supported by the strongest evidence or most aligned with clinical guidelines. Ensure your reasoning supports why this option is the single best choice and how it addresses the clinical situation effectively.

#### A.2 Open-Ended Question Answering Prompt (GPT o1-preview)

You are a cardiologist. Your task is to provide the most specific and concrete reasoning and answer to a general practitioner’s question. Be as specific as possible in your reasoning and answer to the question by providing exact treatment medication, treatment plan, or any other detailed information that is asked about in the question.

**Key Instructions**

1. Answer the question with only one best, most appropriate, and concrete option based on the scenario provided. Avoid giving multiple recommendations or answers, even if more than one seems relevant.
2. Provide reasoning and an explanation for your answer under the section **Reasoning:** to justify your choice.
3. Provide your final answer under the section **Answer:**. This section should contain a single, concise, and definitive answer.

**General Tips**

Be specific and detailed in your reasoning. Jailor your response to the type of question asked, whether it is about a recommended test, treatment plan, medication, or any other action. If multiple options seem equally valid, choose the one supported by the strongest evidence or most aligned with clinical guidelines. Ensure your reasoning supports why this option is the single best choice and how it addresses the clinical situation effectively.

### Section B

To create an automated evaluation pipeline, we used two instances of GPT-4o as evaluators – one for answer evaluation and one for reasoning evaluation. For the open-ended experiments, the answer evaluation agent was provided with the question, the predicted answer, and each answer option in the original multiple-choice question individually and was asked to determine if the predicted answer is entailed by the answer option. If and only if the predicted answer was entailed by the ground truth answer, it was marked correct. For the multiple-choice experiments, the answer correctness of the selected letter (A, B, C, etc.) was determined by string matching to the ground-truth letter. For both open-ended and multiple-choice experiments, the reasoning evaluation agent was provided with the question, the predicted reasoning, and the ground truth reasoning and asked to determine if the predicted reasoning was correct or not. The final output of GPT-4o was a binary “correct/incorrect” for each evaluation. See **Supplementary Section B.1** and **Supplementary Section B.2** for specific prompts used for the evaluation.

#### B.1 Open-ended Answer Evaluation Prompt (GPT-4o)

You are a cardiologist. You will be given a predicted answer and a sample answer to a cardiology question. Your role is to determine if the predicted answer is entailed by the sample answer and why.

#### B.2 Multiple Choice & Open-ended Reasoning Evaluation Prompt (GPT-4o)

You are a cardiologist. You will be provided a cardiology question, the correct answer, a predicted reasoning, and a gold standard ground truth reasoning. Your role is to determine if the predicted reasoning is correct or not based on the gold standard reasoning and provide a justification as to why.

**Key Instructions**

1. Determine if the predicted reasoning is correct based on the gold standard reasoning.
2. Provide a justification for your decision. This should be a detailed explanation of why the predicted reasoning is correct or incorrect based on the gold standard reasoning.

**General Tips**

Ensure the “Ts Correct” field is only “True” or “False.” Be specific and detailed in your justification. Ensure your justification supports why the predicted reasoning is correct or incorrect based on the gold standard reasoning.

### Section C: Agreement between human-expert and GPT-4o evaluators

Across the baseline and modified question experiments, Cohen’s kappa for answer correctness in the baseline open-ended set was 0.37 and 0.54 for reasoning correctness. For baseline multiple-choice questions, the kappa for reasoning correctness was higher at 0.78, representing substantial agreement with the human expert. In the modified open-ended experiment, however, the kappa for answer correctness was only 0.36, suggesting that GPT-4o did not align with expert cardiologists when grading open-ended questions.

## Acknowledgement

We acknowledge Moaz Kamel for his contribution to the initial phase of ACCSAP question data collection.

## Financial Disclosure

Chieh-Ju Chao, MD, is supported by research funding from the CV Prospective award from the Mayo Clinic Department of Cardiovascular Medicine and the AI/ML Enablement award from the Center for Digital Health at the Mayo Clinic. The Stanford team is supported by the Verdant Foundation and Microsoft Azure Cloud Credits.

## Notes

### Competing Interest Statement

The authors have declared no competing interest.

### Summary of Updates

1. Title 2. Corrected format and table/image size to show the cut-off areas in the prior version

## References

1. Nori H, King N, McKinney SM, Carignan D, Horvitz E. Capabilities of GPT-4 on Medical Challenge Problems. arXiv 2023.

2. Omiye JA, Gui H, Rezaei SJ, Zou J, Daneshjou R. Large Language Models in Medicine: The Potentials and Pitfalls: A Narrative Review. Ann Intern Med 2024;177:210–220.

3. OpenAI,:, Hurst A, Lerer A, Goucher AP, Perelman A, Ramesh A, Clark A, Ostrow A, Welihinda A, Hayes A, Radford A, Mądry A, Baker-Whitcomb A, Beutel A, Borzunov A, Carney A, Chow A, Kirillov A, Nichol A, Paino A, Renzin A, Passos AT, Kirillov A, Christakis A, Conneau A, Kamali A, Jabri A, Moyer A, Tam A, Crookes A, Tootoochian A, Tootoonchian A, Kumar A, Vallone A, Karpathy A, Braunstein A, Cann A, Codispoti A, Galu A, Kondrich A, Tulloch A, Mishchenko A, Baek A, Jiang A, Pelisse A, Woodford A, Gosalia A, Dhar A, Pantuliano A, Nayak A, Oliver A, Zoph B, Ghorbani B, Leimberger B, Rossen B, Sokolowsky B, Wang B, Zweig B, Hoover B, Samic B, McGrew B, Spero B, Giertler B, Cheng B, Lightcap B, Walkin B, Quinn B, Guarraci B, Hsu B, Kellogg B, Eastman B, Lugaresi C, Wainwright C, Bassin C, Hudson C, Chu C, Nelson C, Li C, Shern CJ, Conger C, Barette C, Voss C, Ding C, Lu C, Zhang C, Beaumont C, Hallacy C, Koch C, Gibson C, Kim C, Choi C, McLeavey C, Hesse C, Fischer C, Winter C, Czarnecki C, Jarvis C, Wei C, Koumouzelis C, Sherburn D, Kappler D, Levin D, Levy D, Carr D, Farhi D, Mely D, Robinson D, Sasaki D, Jin D, Valladares D, Tsipras D, Li D, Nguyen DP, Findlay D, Oiwoh E, Wong E, Asdar E, Proehl E, Yang E, Antonow E, Kramer E, Peterson E, Sigler E, Wallace E, Brevdo E, Mays E, Khorasani F, Such FP, Raso F, Zhang F, Lohmann F von, Sulit F, Goh G, Oden G, Salmon G, Starace G, Brockman G, Salman H, Bao H, Hu H, Wong H, Wang H, Schmidt H, Whitney H, Jun H, Kirchner H, Pinto HP de O, Ren H, Chang H, Chung HW, Kivlichan I, O’Connell I, O’Connell I, Osband I, Silber I, Sohl I, Okuyucu I, Lan I, Kostrikov I, Sutskever I, Kanitscheider I, Gulrajani I, Coxon J, Menick J, Pachocki J, Aung J, Betker J, Crooks J, Lennon J, Kiros J, Leike J, Park J, Kwon J, Phang J, Teplitz J, Wei J, Wolfe J, Chen J, Harris J, Varavva J, Lee JG, Shieh J, Lin J, Yu J, Weng J, Tang J, Yu J, Jang J, Candela JQ, Beutler J, Landers J, Parish J, Heidecke J, Schulman J, Lachman J, McKay J, Uesato J, Ward J, Kim JW, Huizinga J, Sitkin J, Kraaijeveld J, Gross J, Kaplan J, Snyder J, Achiam J, Jiao J, Lee J, Zhuang J, Harriman J, Fricke K, Hayashi K, Singhal K, Shi K, Karthik K, Wood K, Rimbach K, Hsu K, Nguyen K, Gu-Lemberg K, Button K, Liu K, Howe K, Muthukumar K, Luther K, Ahmad L, Kai L, Itow L, Workman L, Pathak L, Chen L, Jing L, Guy L, Fedus L, Zhou L, Mamitsuka L, Weng L, McCallum L, Held L, Ouyang L, Feuvrier L, Zhang L, Kondraciuk L, Kaiser L, Hewitt L, Metz L, Doshi L, Aflak M, Simens M, Boyd M, Thompson M, Dukhan M, Chen M, Gray M, Hudnall M, Zhang M, Aljubeh M, Litwin M, Zeng M, Johnson M, Shetty M, Gupta M, Shah M, Yatbaz M, Yang MJ, Zhong M, Glaese M, Chen M, Janner M, Lampe M, Petrov M, Wu M, Wang M, Fradin M, Pokrass M, Castro M, Castro MOT de, Pavlov M, Brundage M, Wang M, Khan M, Murati M, Bavarian M, Lin M, Yesildal M, Soto N, Gimelshein N, Cone N, Staudacher N, Summers N, LaFontaine N, Chowdhury N, Ryder N, Stathas N, Turley N, Tezak N, Felix N, Kudige N, Keskar N, Deutsch N, Bundick N, Puckett N, Nachum O, Okelola O, Boiko O, Murk O, Jaffe O, Watkins O, Godement O, Campbell-Moore O, Chao P, McMillan P, Belov P, Su P, Bak P, Bakkum P, Deng P, Dolan P, Hoeschele P, Welinder P, Tillet P, Pronin P, Tillet P, Dhariwal P, Yuan Q, Dias R, Lim R, Arora R, Troll R, Lin R, Lopes RG, Puri R, Miyara R, Leike R, Gaubert R, Zamani R, Wang R, Donnelly R, Honsby R, Smith R, Sahai R, Ramchandani R, Huet R, Carmichael R, Zellers R, Chen R, Chen R, Nigmatullin R, Cheu R, Jain S, Altman S, Schoenholz S, Toizer S, Miserendino S, Agarwal S, Culver S, Ethersmith S, Gray S, Grove S, Metzger S, Hermani S, Jain S, Zhao S, Wu S, Jomoto S, Wu S, Shuaiqi, Xia, Phene S, Papay S, Narayanan S, Coffey S, Lee S, Hall S, Balaji S, Broda T, Stramer T, Xu T, Gogineni T, Christianson T, Sanders T, Patwardhan T, Cunninghman T, Degry T, Dimson T, Raoux T, Shadwell T, Zheng T, Underwood T, Markov T, Sherbakov T, Rubin T, Stasi T, Kaftan T, Heywood T, Peterson T, Walters T, Eloundou T, Qi V, Moeller V, Monaco V, Kuo V, Fomenko V, Chang W, Zheng W, Zhou W, Manassra W, Sheu W, Zaremba W, Patil Y, Qian Y, Kim Y, Cheng Y, Zhang Y, He Y, Zhang Y, Jin Y, Dai Y, Malkov Y. GPT-4o System Card. arXiv 2024.

4. Katz U, Cohen E, Shachar E, Somer J, Fink A, Morse E, Shreiber B, Wolf I. GPT versus Resident Physicians — A Benchmark Based on Official Board Scores. NEJM AI 2024;1.

5. Schubert MC, Wick W, Venkataramani V. Performance of Large Language Models on a Neurology Board–Style Examination. JAMA Netw Open 2023;6:e2346721.

6. Brodeur PG, Buckley TA, Kanjee Z, Goh E, Ling EB, Jain P, Cabral S, Abdulnour R-E, Haimovich A, Freed JA, Olson A, Morgan DJ, Hom J, Gallo R, Horvitz E, Chen J, Manrai AK, Rodman A. Superhuman performance of a large language model on the reasoning tasks of a physician. arXiv 2024.

7. Saab K, Tu T, Weng W-H, Tanno R, Stutz D, Wulczyn E, Zhang F, Strother T, Park C, Vedadi E, Chaves JZ, Hu S-Y, Schaekermann M, Kamath A, Cheng Y, Barrett DGT, Cheung C, Mustafa B, Palepu A, McDuff D, Hou L, Golany T, Liu L, Alayrac J, Houlsby N, Tomasev N, Freyberg J, Lau C, Kemp J, Lai J, Azizi S, Kanada K, Man S, Kulkarni K, Sun R, Shakeri S, He L, Caine B, Webson A, Latysheva N, Johnson M, Mansfield P, Lu J, Rivlin E, Anderson J, Green B, Wong R, Krause J, Shlens J, Dominowska E, Eslami SMA, Chou K, Cui C, Vinyals O, Kavukcuoglu K, Manyika J, Dean J, Hassabis D, Matias Y, Webster D, Barral J, Corrado G, Semturs C, Mahdavi SS, Gottweis J, Karthikesalingam A, Natarajan V. Capabilities of Gemini Models in Medicine. arXiv 2024.

8. Singhal K, Tu T, Gottweis J, Sayres R, Wulczyn E, Amin M, Hou L, Clark K, Pfohl SR, Cole-Lewis H, Neal D, Rashid QM, Schaekermann M, Wang A, Dash D, Chen JH, Shah NH, Lachgar S, Mansfield PA, Prakash S, Green B, Dominowska E, Arcas BA y, Tomašev N, Liu Y, Wong R, Semturs C, Mahdavi SS, Barral JK, Webster DR, Corrado GS, Matias Y, Azizi S, Karthikesalingam A, Natarajan V. Toward expert-level medical question answering with large language models. Nat Med 2025;31:943–950.

9. Zhong T, Liu Z, Pan Y, Zhang Y, Zhou Y, Liang S, Wu Z, Lyu Y, Shu P, Yu X, Cao C, Jiang H, Chen H, Li Y, Chen J, Hu H, Liu Y, Zhao H, Xu S, Dai H, Zhao L, Zhang R, Zhao W, Yang Z, Chen J, Wang P, Ruan W, Wang H, Zhao H, Zhang J, Ren Y, Qin S, Chen T, Li J, Zidan AH, Jahin A, Chen M, Xia S, Holmes J, Zhuang Y, Wang J, Xu B, Xia W, Yu J, Tang K, Yang Y, Sun B, Yang T, Lu G, Wang X, Chai L, Li H, Lu J, Sun L, Zhang X, Ge B, Hu X, Zhang L, Zhou H, Zhang L, Zhang S, Liu N, Jiang B, Kong L, Xiang Z, Ren Y, Liu J, Jiang X, Bao Y, Zhang W, Li X, Li G, Liu W, Shen D, Sikora A, Zhai X, Zhu D, Liu T. Evaluation of OpenAI o1: Opportunities and Challenges of AGI. arXiv 2024.

10. Singhal K, Azizi S, Tu T, Mahdavi SS, Wei J, Chung HW, Scales N, Tanwani A, Cole-Lewis H, Pfohl S, Payne P, Seneviratne M, Gamble P, Kelly C, Babiker A, Schärli N, Chowdhery A, Mansfield P, Demner-Fushman D, Arcas BA y, Webster D, Corrado GS, Matias Y, Chou K, Gottweis J, Tomasev N, Liu Y, Rajkomar A, Barral J, Semturs C, Karthikesalingam A, Natarajan V. Large language models encode clinical knowledge. Nature 2023;620:172–180.

11. Nori H, Lee YT, Zhang S, Carignan D, Edgar R, Fusi N, King N, Larson J, Li Y, Liu W, Luo R, McKinney SM, Ness RO, Poon H, Qin T, Usuyama N, White C, Horvitz E. Can Generalist Foundation Models Outcompete Special-Purpose Tuning? Case Study in Medicine. arXiv 2023.

12. Shahid A, Shetty NS, Patel N, Gaonkar M, Arora G, Arora P. Evaluating Cardiology Certification Using the ACCSAP Question Bank: Large Language Models vs Physicians. Mayo Clin Proc 2025;100:160–163.

13. Lee PC, Sharma SK, Motaganahalli S, Huang A. Evaluating the Clinical Decision-Making Ability of Large Language Models Using MKSAP-19 Cardiology Questions. JACC: Adv 2023;2:100658.

14. Gendler M, Nadkarni GN, Sudri K, Cohen-Shelly M, Glicksberg BS, Efros O, Soffer S, Klang E. Large Language Models in Cardiology: A Systematic Review. medRxiv 2024:2024.09.01.24312887.

15. Mbakwe AB, Lourentzou I, Celi LA, Mechanic OJ, Dagan A. ChatGPT passing USMLE shines a spotlight on the flaws of medical education. PLOS Digit Heal 2023;2:e0000205.

16. Hift RJ. Should essays and other “open-ended”-type questions retain a place in written summative assessment in clinical medicine? BMC Méd Educ 2014;14:249.

17. Tangianu F, Mazzone A, Berti F, Pinna G, Bortolotti I, Colombo F, Nozzoli C, Regina ML, Greco A, Filannino C, Silingardi M, Nardi R. Are multiple-choice questions a good tool for the assessment of clinical competence in Internal Medicine? Ital J Med 2018;12:88–96.

18. Wijk EV van, Janse RJ, Ruijter BN, Rohling JHT, Kraan J van der, Crobach S, Jonge M de, Beaufort AJ de, Dekker FW, Langers AMJ. Use of very short answer questions compared to multiple choice questions in undergraduate medical students: An external validation study. PLOS ONE 2023;18:e0288558.

19. Parekh P, Bahadoor V. The Utility of Multiple-Choice Assessment in Current Medical Education: A Critical Review. Cureus 2024;16:e59778.

20. Hosseini P, Sin JM, Ren B, Thomas BG, Nouri E, Farahanchi A, Hassanpour S. A Benchmark for Long-Form Medical Question Answering. arXiv 2024.

21. Brush JE, Sherbino J, Norman GR. Diagnostic reasoning in cardiovascular medicine. BMJ 2022;376:e064389.

22. Kim J, Podlasek A, Shidara K, Liu F, Alaa A, Bernardo D. Limitations of Large Language Models in Clinical Problem-Solving Arising from Inflexible Reasoning. arXiv 2025.

23. OpenAI,:, Jaech A, Kalai A, Lerer A, Richardson A, El-Kishky A, Low A, Helyar A, Madry A, Beutel A, Carney A, Iftimie A, Karpenko A, Passos AT, Neitz A, Prokofiev A, Wei A, Tam A, Bennett A, Kumar A, Saraiva A, Vallone A, Duberstein A, Kondrich A, Mishchenko A, Applebaum A, Jiang A, Nair A, Zoph B, Ghorbani B, Rossen B, Sokolowsky B, Barak B, McGrew B, Minaiev B, Hao B, Baker B, Houghton B, McKinzie B, Eastman B, Lugaresi C, Bassin C, Hudson C, Li CM, Bourcy C de, Voss C, Shen C, Zhang C, Koch C, Orsinger C, Hesse C, Fischer C, Chan C, Roberts D, Kappler D, Levy D, Selsam D, Dohan D, Farhi D, Mely D, Robinson D, Tsipras D, Li D, Oprica D, Freeman E, Zhang E, Wong E, Proehl E, Cheung E, Mitchell E, Wallace E, Ritter E, Mays E, Wang F, Such FP, Raso F, Leoni F, Tsimpourlas F, Song F, Lohmann F von, Sulit F, Salmon G, Parascandolo G, Chabot G, Zhao G, Brockman G, Leclerc G, Salman H, Bao H, Sheng H, Andrin H, Bagherinezhad H, Ren H, Lightman H, Chung HW, Kivlichan I, O’Connell I, Osband I, Gilaberte IC, Akkaya I, Kostrikov I, Sutskever I, Kofman I, Pachocki J, Lennon J, Wei J, Harb J, Twore J, Feng J, Yu J, Weng J, Tang J, Yu J, Candela JQ, Palermo J, Parish J, Heidecke J, Hallman J, Rizzo J, Gordon J, Uesato J, Ward J, Huizinga J, Wang J, Chen K, Xiao K, Singhal K, Nguyen K, Cobbe K, Shi K, Wood K, Rimbach K, Gu-Lemberg K, Liu K, Lu K, Stone K, Yu K, Ahmad L, Yang L, Liu L, Maksin L, Ho L, Fedus L, Weng L, Li L, McCallum L, Held L, Kuhn L, Kondraciuk L, Kaiser L, Metz L, Boyd M, Trebacz M, Joglekar M, Chen M, Tintor M, Meyer M, Jones M, Kaufer M, Schwarzer M, Shah M, Yatbaz M, Guan MY, Xu M, Yan M, Glaese M, Chen M, Lampe M, Malek M, Wang M, Fradin M, McClay M, Pavlov M, Wang M, Wang M, Murati M, Bavarian M, Rohaninejad M, McAleese N, Chowdhury N, Chowdhury N, Ryder N, Tezak N, Brown N, Nachum O, Boiko O, Murk O, Watkins O, Chao P, Ashbourne P, Izmailov P, Zhokhov P, Dias R, Arora R, Lin R, Lopes RG, Gaon R, Miyara R, Leike R, Hwang R, Garg R, Brown R, James R, Shu R, Cheu R, Greene R, Jain S, Altman S, Toizer S, Toyer S, Miserendino S, Agarwal S, Hernandez S, Baker S, McKinney S, Yan S, Zhao S, Hu S, Santurkar S, Chaudhuri SR, Zhang S, Fu S, Papay S, Lin S, Balaji S, Sanjeev S, Sidor S, Broda T, Clark A, Wang T, Gordon T, Sanders T, Patwardhan T, Sottiaux T, Degry T, Dimson T, Zheng T, Garipov T, Stasi T, Bansal T, Creech T, Peterson T, Eloundou T, Qi V, Kosaraju V, Monaco V, Pong V, Fomenko V, Zheng W, Zhou W, McCabe W, Zaremba W, Dubois Y, Lu Y, Chen Y, Cha Y, Bai Y, He Y, Zhang Y, Wang Y, Shao Z, Li Z. OpenAI o1 System Card. arXiv 2024.

24. Otto CM, Nishimura RA, Bonow RO, Carabello BA, Erwin JP, Gentile F, Jneid H, Krieger EV, Mack M, McLeod C, O’Gara PT, Rigolin VH, Sundt TM, Thompson A, Toly C. 2020 ACC/AHA Guideline for the Management of Patients With Valvular Heart Disease A Report of the American College of Cardiology/American Heart Association Joint Committee on Clinical Practice Guidelines. J Am Coll Cardiol 2020.

25. Stout KK, Daniels CJ, Aboulhosn JA, Bozkurt B, Broberg CS, Colman JM, Crumb SR, Dearani JA, Fuller S, Gurvitz M, Khairy P, Landzberg MJ, Saidi A, Valente AM, Hare GFV. 2018 AHA/ACC Guideline for the Management of Adults With Congenital Heart Disease: A Report of the American College of Cardiology/American Heart Association Task Force on Clinical Practice Guidelines. J Am Coll Cardiol 2019;73:e81 e192.

26. Ommen SR, Mital S, Burke MA, Day SM, Deswal A, Elliott P, Evanovich LL, Hung J, Joglar JA, Kantor P, Kimmelstiel C, Kittleson M, Link MS, Maron MS, Martinez MW, Miyake CY, Schaff HV, Semsarian C, Sorajja P, Hung J. 2020 AHA/ACC Guideline for the Diagnosis and Treatment of Patients With Hypertrophic Cardiomyopathy. Circulation 2020;142:e558–e631.

27. Builoff V, Shanbhag A, Miller RJH, Dey D, Liang JX, Flood K, Bourque JM, Chareonthaitawee P, Phillips LM, Slomka PJ. Evaluating AI proficiency in nuclear cardiology: Large language models take on the board preparation exam. J Nucl Cardiol 2025;45:102089.

28. Huwiler J, Oechslin L, Biaggi P, Tanner FC, Wyss CA. Experimental assessment of the performance of artificial intelligence in solving multiple-choice board exams in cardiology. Swiss Méd Wkly 2024;154:3547.

29. Milutinovic S, Petrovic M, Begosh-Mayne D, Lopez-Mattei J, Chazal RA, Wood MJ, Escarcega RO. Evaluating Performance of ChatGPT on MKSAP Cardiology Board Review Questions. Int J Cardiol 2024;417:132576.

30. Altamimi I, Alhumimidi A, Alshehri S, Alrumayan A, Al-khlaiwi T, Meo SA, Temsah M-H. The scientific knowledge of three large language models in cardiology: multiple-choice questions examination-based performance. Ann Med Surg 2024;86:3261–3266.

31. Gupta V, Pantoja D, Ross C, Williams A, Ung M. Changing Answer Order Can Decrease MMLU Accuracy. arXiv 2024.

32. Wu S, Peng Z, Du X, Zheng T, Liu M, Wu J, Ma J, Li Y, Yang J, Zhou W, Lin Q, Zhao J, Zhang Z, Huang W, Zhang G, Lin C, Liu JH. A Comparative Study on Reasoning Patterns of OpenAI’s o1 Model. arXiv 2024.

33. Hager P, Jungmann F, Holland R, Bhagat K, Hubrecht I, Knauer M, Vielhauer J, Makowski M, Braren R, Kaissis G, Rueckert D. Evaluation and mitigation of the limitations of large language models in clinical decision-making. Nat Med 2024;30:2613–2622.

34. Reese JT, Danis D, Caufield JH, Groza T, Casiraghi E, Valentini G, Mungall CJ, Robinson PN. On the limitations of large language models in clinical diagnosis. medRxiv 2024:2023.07.13.23292613.

35. Groves M, O’Rourke P, Alexander H. The clinical reasoning characteristics of diagnostic experts. Méd Teach 2003;25:308–313.

36. Bavaresco A, Bernardi R, Bertolazzi L, Elliott D, Fernández R, Gatt A, Ghaleb E, Giulianelli M, Hanna M, Koller A, Martins AFT, Mondorf P, Neplenbroek V, Pezzelle S, Plank B, Schlangen D, Suglia A, Surikuchi AK, Takmaz E, Testoni A. LLMs instead of Human Judges? A Large Scale Empirical Study across 20 NLP Evaluation Tasks. arXiv 2024.

37. Szymanski A, Ziems N, Eicher-Miller HA, Li TJ-J, Jiang M, Metoyer RA. Limitations of the LLM-as-a-Judge Approach for Evaluating LLM Outputs in Expert Knowledge Tasks. Proc 30th Int Conf Intell User Interfaces 2025:952–966.

